# An Age and Space Structured SIR Model Describing the Covid-19 Pandemic

**DOI:** 10.1101/2020.05.15.20103317

**Authors:** R.M. Colombo, M. Garavello, F. Marcellini, E. Rossi

**Affiliations:** INdAM Unit, University of Brescia, via Branze, 38, 25123 Brescia, Italy; Università degli Studi di Milano Bicocca, Dipartimento di Matematica e Applicazioni, via R. Cozzi, 55, 20126 Milano, Italy

**Keywords:** Age and Space Structured SIR Model, Differential Equations in Epidemic Modeling, Covid-19 Modeling

## Abstract

We present an epidemic model capable of describing key features of the present Covid-19 pandemic. While capturing several qualitative properties of the virus spreading, it allows to compute the basic reproduction number, the number of deaths due to the virus and various other statistics. Numerical integrations are used to illustrate the relevance of quarantine and the role of care houses.

## 1 Introduction

Our aim here is to present a model that contains key features of the Covid-19 outbreak. It uses as starting point the classical SIR class of models, see [19, § 13.5] but is thoroughly adapted to the present day pandemic. Indeed, its key features are:

- Infected individuals are distinguished between Infective (*I*) and Hospitalized (*H*). The former ones do spread the disease, while the latter ones, hospitalized or in quarantine, don’t. We thus consider the four populations of Susceptible (*S*), Infective (*I*), Hospitalized (*H*) and Recovered (*R*) individuals.
- The four densities *S, I, H, R* depend on time *t* ∈ ℝ_+_, on age *a* ∈ ℝ_+_ and on a space coordinate *x* ∈ ℝ^2^. *S*(*t, a, x*) (respectively *I*(*t, a, x*), *H*(*t, a, x*), *R*(*t, a, x*)) quantifies the individuals of type *S* (respectively *I, H, R*) that at time *t* are of age *a* and are sited at position *x*.
- Infection is propagated in space: *S* individuals can be infected by *I* individuals of all ages, provided they are at the same time at a distance less than a given threshold. *H* individuals do not infect anyone.
- *S, I* and *R* individuals move in the space domain with a time, age and space dependent velocity. *H* individuals are not assumed to move. A further distinction of *S* (respectively *I* and *R*) individuals according to their different destinations is also possible.
- At a given time, age and space dependent rate, infective individuals (*I*) are hospitalized or constrained to quarantine, thus entering the *H* population. Both infective and hospitalized individuals recover or die at time, age and space dependent rates.

Before passing to the rigorous description of the model, we recall that different countries reacted to Covid-19 in different ways. Nevertheless, the initial stages of the virus spreading are quantitatively quite similar in the different countries, once they are correctly scaled with respect to the overall population: their only difference is essentially a time delay, see Figure 1. These striking similarities definitely justify the search for a unique model able to describe the initial virus spreading.

**Figure 1:**
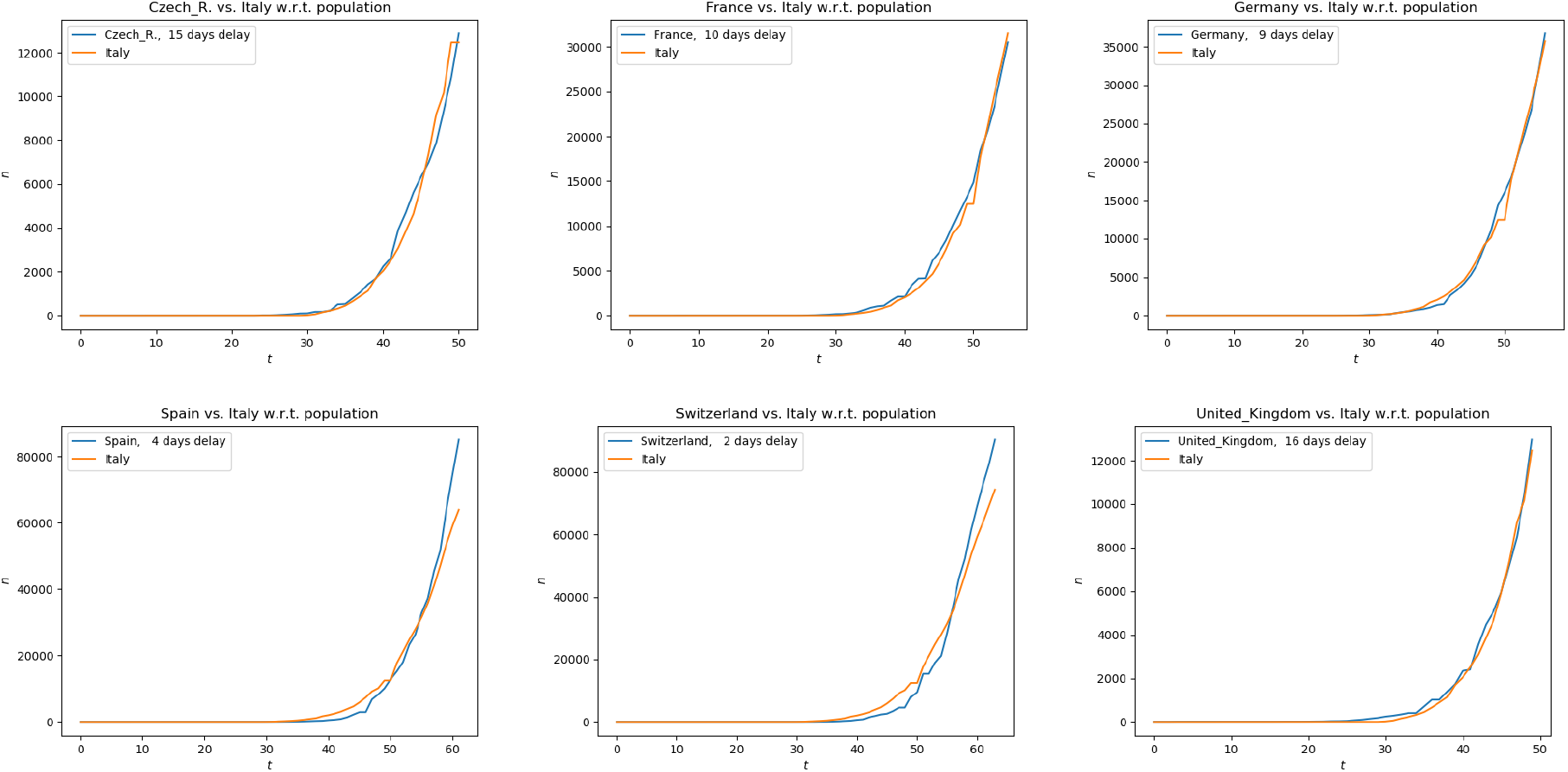
Number of confirmed Covid-19 cases in different European countries, scaled to the Italian population, as a function of time. Along each horizontal axis, time is measured in days starting January 22nd, 2020, each country compared to Italy being delayed by the amount indicated in each top left corner. The numbers of cases are taken from [11], the populations’ data from [22].

As is well known, different countries are taking different measures to contrast the pandemic. Key differences typically concern the strength of *lock down*, constraining individuals’ movements and contacts at different levels. In the model presented below, these differences can be covered through *ad hoc* choices of a function, namely *ρ*, that can describe various types of contagion. We describe below some effects of care houses, that is of places where the virus spreads faster, in accelerating the infection. As a further example, we show the effects of different quarantine policies, only from the pandemic evolution point of view. In this connection, we note the possible interest in extending the model presented below to possibly cover also some of the consequences of the pandemic at the economic/industrial/financial levels.

Below, we use the model here introduced to describe qualitatively relevant features of the Covid-19 pandemic. At a quantitative level, the use of the present model relies on the availability of reliable data, which is not always possible. In this connection, we refer for instance to [3] for the description of a method able to cope with uncertain data.

For completeness, we refer to [16] for a different approach to the modeling of the Covid-19 pandemic, also based on integro–differential equations. Differently from the model therein, here we do not resort to delayed terms, partly using the standard coupling between the different equations describing the passage between different populations. A related work considering an epidemic model with age structure and immigration is described in [13].

The next section is devoted to a more rigorous formulation of the model, of its key approximations and of its main properties. Then, by means of numerical integrations, we show key qualitative properties of the solutions, which agree with well known properties of the Covid-19 pandemic. In these integrations, the various functions entering the model definitions are chosen in agreement with publicly available data.

## 2 The Model

A population lives in a region χ ⊆ ℝ*^n^* and is subject to an infective disease. Clearly, we typically set *n* = 2, but also the case *n* = 1 can be of use in a simplified framework.

Throughout, *S* = *S*(*t, a, x*) is the number of susceptible individuals at time *t* ∈ ℝ_+_, of age *a* ∈ ℝ_+_ at position *x* ∈ χ. When infected, susceptible individuals enter the *I* population, i.e., they first turn into being infected and infective, possibly asymptomatic. These individuals are then hospitalized or set into quarantine at a rate *κ* = *κ*(*t, a, x*) and, when this happens, we label them as *H* = *H*(*t, a, x*). Both *I*, respectively *H*, individuals may possibly recover at rates *ϑ* = *ϑ*(*t, a, x*), respectively *η* = *η*(*t, a, x*), entering the population labeled as *R* = *R*(*t, a, x*). We keep the *R* population distinct from the *S* one, assuming that those who recover are immune to any further infection. A different assumption, namely that those who recover are not immune, amounts, for instance, to add further terms coupling the last equation to the previous ones in (2.1).

*S* (respectively *I* and *R*) individuals move in space with the assigned velocity *v_S_* = *v_S_*(*t, a, x*) (respectively *v_I_* = *v_I_*(*t, a, x*) and *v_R_* = *v_R_*(*t, a, x*)). Depending on the geographical scale at which the present model is applied, it might be of use the more general dynamics suggested by crowd dynamics model, see for instance [7]. At a different scale, *v_S_, v_I_* and *v_R_* may also describe the collective movements of relatively large sets of individuals heading towards regions less hit by the pandemic.

Independently of the movements’ scale, when individuals of the same type, say *S*, follow different routes, we distinguish *S* into different components, say *S*^1^, *S*^2^, *…*, and we assign them the different velocities *v_S_*^1^, *v_S_*^2^, *…*, following a usual approach in crowd dynamics, see for instance [9]. However, this latter distinction introduces a non trivial formal complexity, with no relevance at the level of the present initial description and we leave the corresponding technical details to a later work.

The disease is transmitted by *I* individuals to *S* ones that are, at any given time, geographically near, independently of age.

We are thus lead to the model

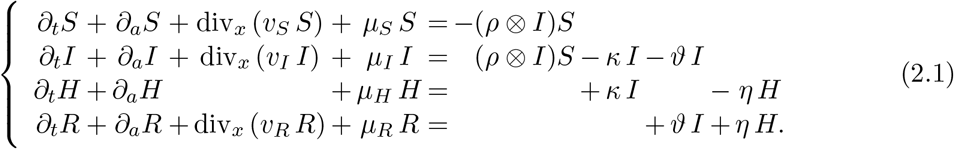

Here, the term *κ I* describes the speed at which *I* individuals are hospitalized or put into quarantine. Similarly, the term *ϑ I* is the speed at which *I* individuals recover, while hospitalized individuals recover with a rate *η H*. As usual, for *A* = *S, I, H, R, µ_A_* is the mortality of the individuals of type *A*. All the above parameters, in particular the mortality rates, are time, age and space dependent.

In (2.1), for merely typographical reasons, we use the abbreviation

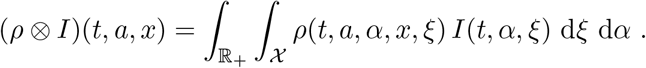

This key term is the rate at which susceptible individuals get infected. The function *ρ* plays a fundamental role, for it describes the dynamics of the disease transmission. Various properties of the function *ρ* have a clear counterpart on the real characteristics of the virus spreading. Depending on the particular scenario that is under consideration, different choices of *ρ* are due. However, the following key property is essential:

### Virus Transmission

Assume that *δ* is the smallest distance satisfying

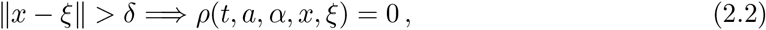

*x* and *ξ* being positions in χ. Then, *δ* represents the maximal distance at which the virus can be transmitted. Note that the speed of the infection is infinite, within the distance *δ*.

Remark that suitable choices of *ρ* may well describe various specific situations. For instance, the dependence of *ρ* on the age variables *a* and *α* allows to consider situations in which contagion is restricted – or more/less prominent – among individuals of specific ages, for instance of the same age.

Model (2.1) needs to be complemented with initial and boundary data, say

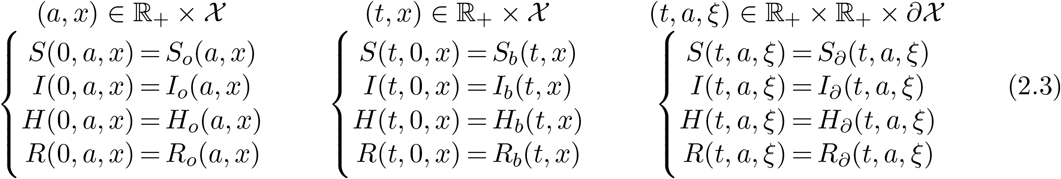

which have to be chosen according to the specific situation under study. The only general constraint to be imposed on these data, besides obvious minimal regularity conditions necessary from the analytic point of view, is that newborns, corresponding to *a* = 0, are mostly in the *S* population. In other words, while the analytic well posedness is completely independent of this requirement, we expect that in every realistic application we have

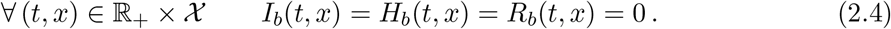

As it is well known, in the case of general balance laws, assigning and understanding the role of the boundary condition along the spatial boundary *∂*X requires particular care, see [4,20,21]. Here, though not strictly necessary form the analytic point of view, we assume the individuals’ velocities to be assigned are time, age and space dependent functions, so that boundary data are essential whenever the velocities point inward X, while they are neglected when velocities point outward.

A relevant time dependent statistics commonly used to quantify the spreading speed of the disease is the basic reproduction number [18, Section 10.2], typically denoted by ℛ_*o*_:

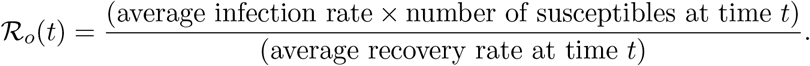

Above *“average”* refers to both age and space averages. In the present *dynamic* setting, this index needs to be time dependent. Moreover, the presence of 2 different populations of ill individuals, namely the infective (*I*) and the hospitalized (*H*) ones, allows for the introduction of two indexes inspired by ℛ_*o*_. The first one, say ℛ_*o*_, considers only the infective ones while the latter, say Q_*o*_, comprises also the hospitalized ones:

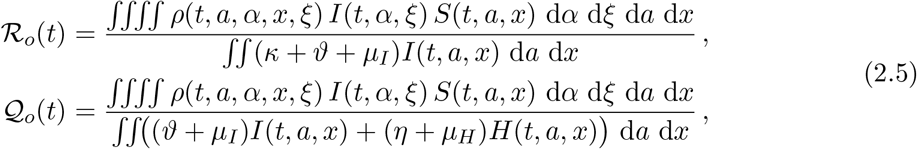

where we shortened *κ* = *κ*(*t, a, x*), *ϑ* = *ϑ*(*t, a, x*), *η* = *η*(*t, a, x*), *µ_I_* = *µ_I_*(*t, a, x*) and *µ_H_* = *µ_H_*(*t, a, x*).

The above definitions are justified by the following necessary and sufficient conditions, that hold provided the inflow/outflow in/from *χ* vanishes and provided no newborn is ill:

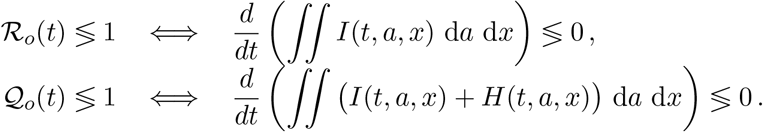

The proofs amount to elementary applications of the Divergence Theorem and, hence, are omitted.

In other words, ℛ_*o*_(*t*) describes the instantaneous variation of the number of infective (*I*) individuals at time *t*, while 𝒬_*o*_(*t*) describes that of the total number of ill (*I* + *H*) individuals. Thus, ℛ_0_ measures the danger of being infected, while Q_0_ measures the overall effect of the disease spreading, coherently with (2.2).

For completeness, we note that the above definitions can be slightly simplified neglecting the mortality terms, obtaining

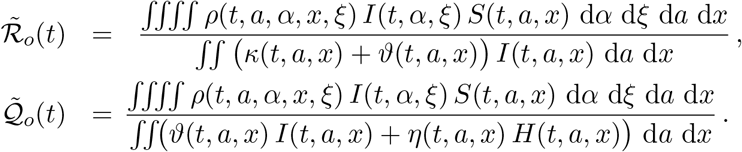

These latter simplified expressions still give useful information, since

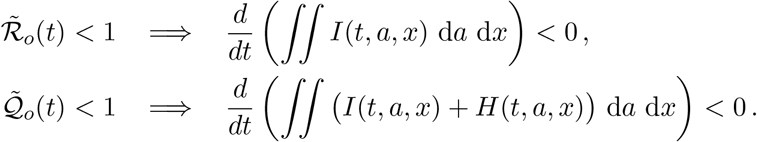

We remark that on relatively short time intervals (up to, say, a year or so), the difference between ℛ_0_ and 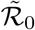 (or between 𝒬_0_ and 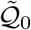) is likely to be negligible.

As a further remark, note that under assumption (2.4), the instantaneous variation in the total population in the region χ is

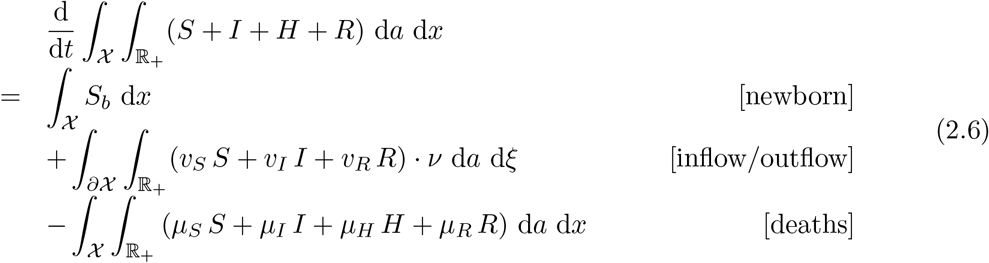

where *ν* = *ν*(*ξ*) is the inward normal at *ξ* to *∂*χ and the boundary data *S_b_* measures newborns, see (2.3). The equality (2.6) clearly shows the role of the mortality rates *µ_S_, µ_I_, µ_H_, µ_R_*.

An obvious consequence of (2.1) is that, for the epidemic to arise, it is necessary that infective individuals are either present or enter the domain χ. Indeed, if *I_o_*(*a, x*) ≡ 0 and *I_∂_*(*t, a, ξ*) ≡ 0, then the whole population remains forever untouched by the virus.

A key structural property of (2.1) is that the first two equations are independent of the latter two. Once *S* and *I* are known, the explicit forms of *H* and *R* are available through, for instance, a mixing of [10, Lemma 4.10] and [6, Lemma 3]. Formally, system (2.1) is a system of balance laws in several space dimensions. For these kind of partial differential equations, a general well posedness theory is still missing. However, the different equations are coupled through the source terms, similarly to the cases considered in [9, 8] where well posedness is obtained, as well as the stability with respect to the parameters defining the equation, see [17].

Different *costs* are related to the pandemic. First, the total number of deaths due to the disease on the time interval [0, *T*], say 𝒟(*T*), is probably the most relevant one:

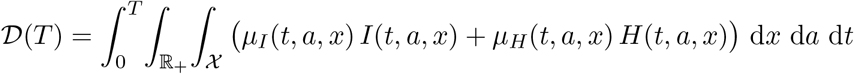

We do not enter here the issue of assigning the cause of the death to the virus in presence of other health problems.

On the other hand, we can also consider a more general cost comprising, for instance, also the expenses that the health system must sustain. Therefore, we refer to the cost functional

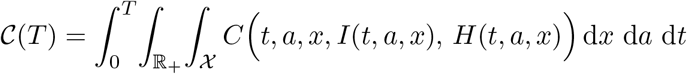

Above, the explicit dependence of *C* on *t, a, x* may account for the peculiarities that different time periods, ages or regions may have.

The many policies or strategies that can be adopted to confine the infection enter *ρ* and the various parameters in (2.1). Besides, a quite natural choice is to use as control the function *κ*, since it describes the rate at which infective individuals are confined.

Before passing to simplified versions of (2.1), we note that generalizations and extensions are also possible. First, each population can be split into females and males, for instance. On long time intervals, the introduction of *growth functions*, accounting for the different aging of the different populations, might also be considered.

Hopefully, a particularly hot topic in the next future will be the strategy to adopt when a vaccine will be available. From the modeling point of view, this amounts to insert vaccination in the present model, following the framework in [5].

Finally we recall that, in system (2.1), recovered individuals can not be infected again. The mostly unfortunate case where this assumption were false would amount to the introduction of further terms on the right hand sides.

## 2.1 Simplified Versions

While the model (2.1) looks quite general, in its use on a time scale of, say, a year or less, the terms *∂_a_S, ∂_a_I, ∂_a_H* and *∂_a_R*, typically describing the aging of the population, can be neglected. Then, (2.1) reduces to the system of partial differential equations

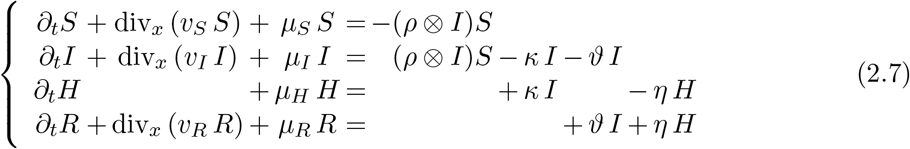

where the age variable *a* plays the role of a parameter. Note that, as in the general case, the latter two equations can be explicitly solved, as soon as a solution to the system consisting of the former two equations is available.

If moreover we neglect the spatial velocity, i.e, we set *v_S_* = *v_I_* = *v_R_* = 0, then system (2.7) becomes

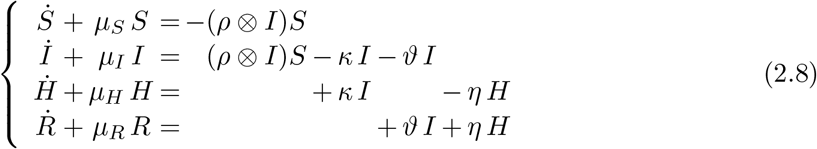

where the dot symbol stands, as usual, for the time derivative. In the latter system, also the space variable *x* plays the role of a parameter.

## 3 Qualitative Properties

The present work aims at a qualitative description of the model (2.1), mainly in its simplified form (2.8). Nevertheless, we first partially justify the choice of the key parameters used in the integrations below. Then, we describe the numerical algorithm used and finally we dive into a few realistic situations seen through the model (2.8).

### 3.1 Parameters’ Choices

We detail here the procedures adopted to select the values of the parameters used.

In the integrations below we are inspired by average Italian data, scaled to the square [−50, 50] × [−50, 50], with a total population of about 2 millions inhabitants, approximating the average Italian population density of 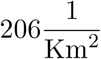.

As common experience shows, age plays a role in the evolution and consequences of the Covid-19 infection. Therefore, we distinguish below 4 age classes (in years): [0, 40], [40, 60],[60, 80[ and [80, +∞[. We recall that the total population is distributed among these age classes, following [14], with the coefficients:

| age class | $[0, 40[$ | $[40, 60[$ | $[60, 80[$ | $[80, +\infty[$ |
| --- | --- | --- | --- | --- |
| density distribution | 0.404 | 0.308 | 0.218 | 0.0700 |

The estimation of all related to the *I* population is intrinsically quite difficult. These individuals are infective but not isolated, therefore they may be unknown to any agency, as they may well be unaware of their status. It is generally believed that their number exceeds the official number of positive tests, but very little seems to be known about the other parameters specific to the *I* population, such as mortality, for instance.

#### 3.1.1 Mortalities

Remark that here our aim is to capture qualitative features or compare different strategies to cope with the pandemic, rather than obtain quantitatively correct forecasts. Therefore, we are more interested in the *ratios* among the different mortalities, rather than in their absolute values. Dimensionally, mortalities are measured by 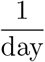.

| Age class | $[0, 40[$ | $[40, 60[$ | $[60, 80[$ | $[80, +\infty[$ | See |
| --- | --- | --- | --- | --- | --- |
| Total Deaths in 2018 | 7893 | 38896 | 182282 | 404062 | [15] |
| Residents in 2018 | 24421783 | 18640004 | 13215186 | 4207000 | [14] |
| $\mu_S(a) = \mu_R(a) =$ | 8.85e-7 | 5.72e-6 | 3.78e-5 | 2.63e-4 | (3.1) |
| Covid-19 deaths (up to 15.03): | 4 | 55 | 746 | 892 | [1] |
| Covid-19 deaths (up to 22.03): | 12 | 209 | 2309 | 2488 | [2] |
| Covid-19 cases (up to 15.03): | 2953 | 7729 | 9561 | 4636 | [1] |
| Covid-19 cases (up to 22.03): | 6891 | 18547 | 21743 | 10514 | [2] |
| $\mu_H(a) = \mu_i(a) =$ | 2.90e-4 | 2.03e-3 | 1.83e-2 | 3.87e-2 | (3.2) |

The mortalities *µ_S_* and *µ_R_* are computed, in each age interval, so that

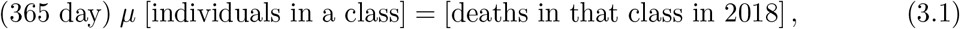

while to compute the values of *µ_H_* we used

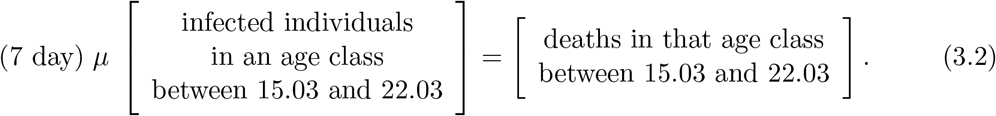

In the above computations leading to the estimates of the mortalities, we assumed that all those who are aware of being infective belong to the *H* population, meaning that they do not infect anyone.

Here, we set *µ_I_* = *µ_H_*. Indeed, the *I* population both comprises asymptomatic individuals that hardly realize being infected, as well as those who are not taken care of. In the average, we arbitrarily assume that their mortality is as big as that of the isolated individuals.

#### 3.1.2 Transitions between Populations

The parameter *κ* = *κ*(*t, a, x*) measures the rate at which infective individuals are *“blocked”*, i.e., hospitalized or set to quarantine. Due to the above recalled nature of the *I* population, we estimate *κ* as being generally larger than *µ_I_*. Indeed, we expect that infective individuals usually become known and, hence, isolated well before their conditions get too bad. Moreover, we expect that *κ* is bigger at higher ages, since the presence of infective individuals that are not aware of their status (and, hence, are not isolated) might be larger at lower ages. Thus, we choose *κ* as being only age dependent and, more precisely, we set^1^

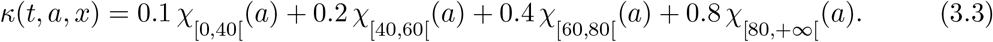

Recall that *κ* plays a key role in the control of the epidemic and a paragraph below is devoted to show its relevance.

The parameter *ϑ* is the speed at which infective individuals recover. It is realistic to assume that this happens at a rate faster than the death rate and, what is more relevant, faster for younger individuals. Thus, neglecting the dependence on time and space, we set

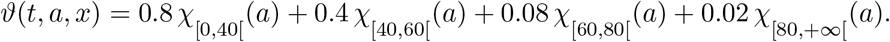

Finally, *η* is the speed at which hospitalized individuals recover. We get from [12] the total number of individuals that recovered on March 23rd, namely 7342 out of a total *H* population on that day of 63927, so that we set

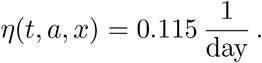

Note that the parameter *η* correctly turns out to be an average of the values attained by *ϑ*.

### 3.2 The Numerical Algorithm

The numerical integration of (2.8) amounts to the approximate solution of ordinary differential equations where age (*a*) and space (*x*) play the role of parameters. The integral coupling the first two equations in the right hand side of (2.8) is computed by means of a quadrature formula. Then, it is added to the other terms in the right hand side of (2.8) and an approximate solution is obtained using the exact solution to the linear ODE consisting of the left hand sides alone. This stratagem allows to comply with the global balance (2.6) of all the populations.

For the sake of completeness, we specify that all meshes are fixed and uniform. The age boundary *a* = 0 as well as the geographic boundary in the *x* variable need no specific treatment, due to the absence of any convective term.

### 3.3 The Role of Quarantine

We now show that the presented model, though in the simplified form (2.8), does capture the relevance of the quarantine. We integrate three instances of (2.8) differing exclusively in the values attained by *κ*. Recall that this parameter quantifies the speed at which infective individuals are confined to quarantine.

In the first case, we set *κ* ≡ 0, then we use *κ* as defined in (3.3), in the third integration we use 10 times the value of *κ* in (3.3) and in the latter integration we use 20 times the *κ* in (3.3).

We detail the choices of the initial datum (2.3):

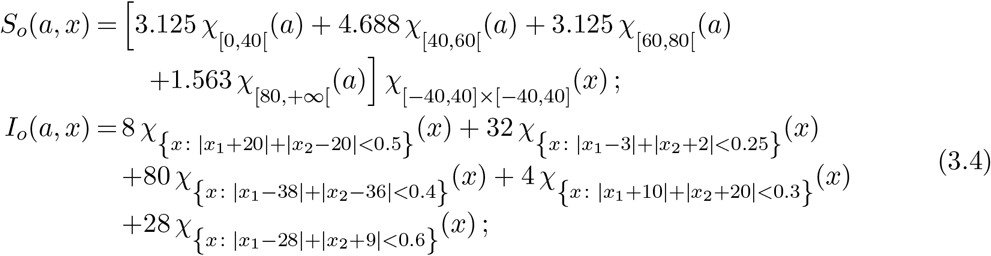

The dynamics of infection is described by the function *ρ* which we here select as follows:

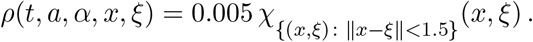

This choice amounts to allow infection to pass from infective to susceptibles only provided that individuals are less than 1.5 apart. The transmission of the disease takes place independently of the age and of the absolute positions, the only constraint being the vicinity of infective and susceptibles. We also choose that the transmission of the disease is independent of time.

The effect of quarantine is well captured even by the simplified model (2.8). When *κ* is 0, no quarantine occurs and the virus spreads the fastest, see Figure 2. Higher values of *κ* mean that more individuals are quarantined/hospitalized, slowing down the spreading of the virus.

**Figure 2:**
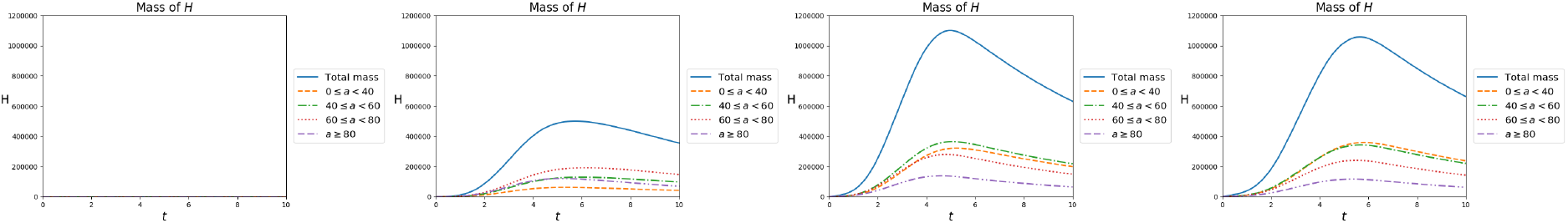
Total number of isolated individuals as a function of time, according to (2.8), in the four cases, from left to right: *κ* = 0, *κ* as in (3.3), 10 *κ* and 20 *κ*. Remarkably, in the latter case the peak of the map *t* → ∬ *H* d*a* d*x* is lower than on the preceding case. Indeed, a high value of *κ* reduces the number of infectives and, as a consequence, may also reduce the total number of individuals in quarantine.

As *κ* increases, *S* individuals take more time to get infected, see Figure 3. Clearly, with lower values of *κ*, the disease spreads more rapidly, so that the number of infectives is far higher, see Figure 4, and less individuals recover, see Figure 5. Moreover the total number of deaths decreases as *κ* increases, see Figure 6.

**Figure 3:**
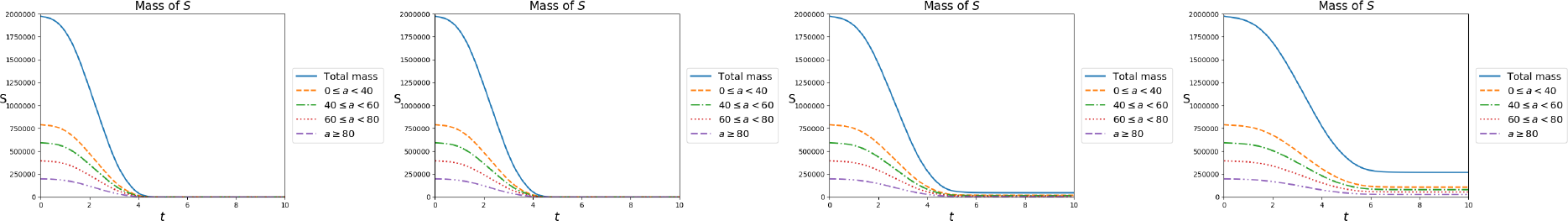
Total number of susceptible individuals as a function of time, according to (2.8), in the four cases, from left to right: *κ* = 0, *κ* as in (3.3), 10 *κ* and 20 *κ*. The increase in the infectives’ isolation speed slightly lengthen the time necessary for susceptibles to be infected.

**Figure 4:**
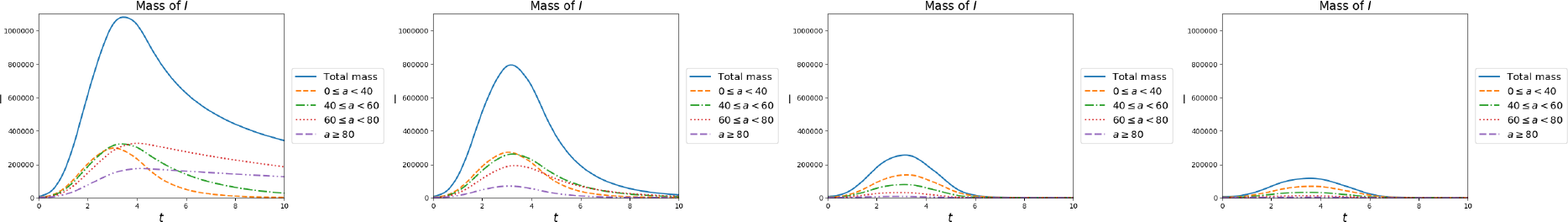
Total number of infective individuals as a function of time, according to (2.8), in the four cases, from left to right: *κ* = 0, *κ* as in (3.3), 10 *κ* and 20 *κ*. It is evident that quarantine sharply reduces the amount of infective individuals.

**Figure 5:**
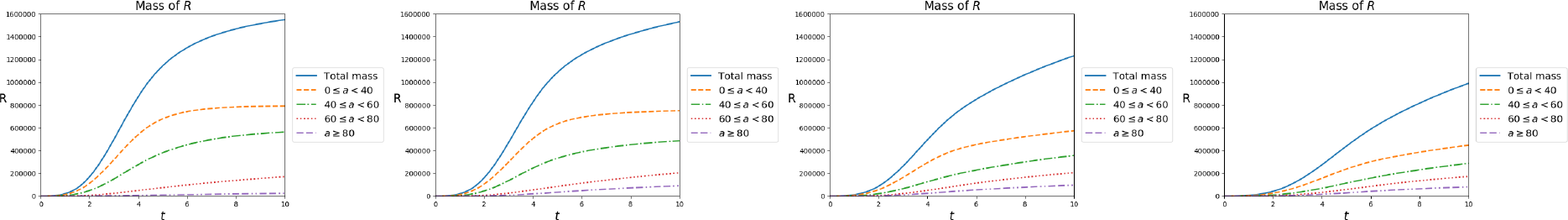
Total number of recovered individuals as a function of time, according to (2.8), in the four cases, from left to right: *κ* = 0, *κ* as in (3.3), 10 *κ* and 20 *κ*. The increase in the quarantined individuals leads to a decrease of the infected ones and, hence, also of those that recover.

**Figure 6:**
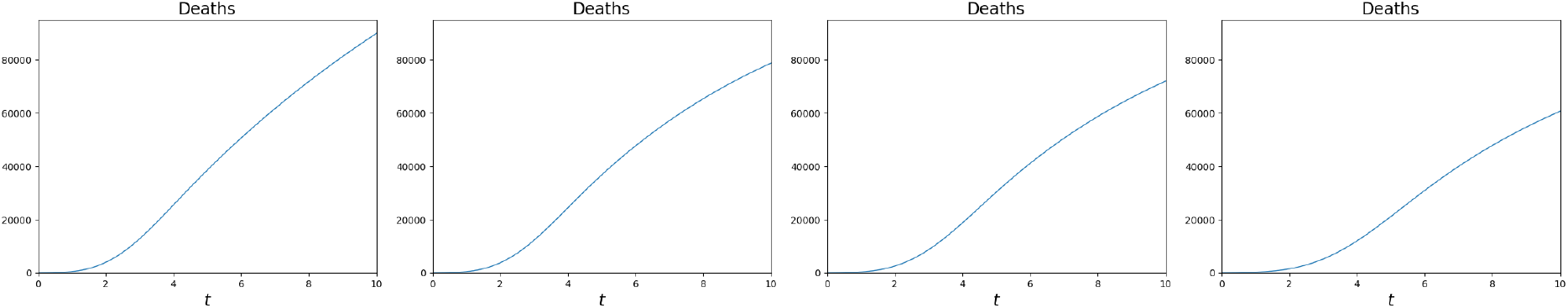
Total number of deaths due to the pandemic as a function of time, according to (2.8), in the four cases, from left to right: *κ* = 0, *κ* as in (3.3), 10 *κ* and 20 *κ*. Here, the effect of quarantine is evident, sharply reducing the death toll.

Note the counter-intuitive effect according to which the highest value of *κ* does not correspond the highest peak in the map *t* → ∬ *H* d*a* d*x*. Indeed, higher values of *κ* reduce the total number of infected people which, as a consequence, may well lead to a *reduction* in the number of isolated individuals, see Figure 2.

Note that higher values of *κ* not only lead to lower values of *t* → ∬ *I*(*t, a, x*) d*a* d*x*, but also move the peak of this function to the right. From the practical point of view, this is likely to correspond to a lighter exploitation of intensive care units, a key aspect from the public health point of view.

This *“slowing”* effect is evident also in Figure 7: lower values of *κ* result in a shorter time interval where ℛ_0_ exceeds 1. However, the values attained by this index may cause an excessive stress on intensive care units. On the contrary, higher values of *κ* lead to a longer period where ℛ_0_ exceeds 1, but with values that suggest a minor stress on the health system.

**Figure 7:**
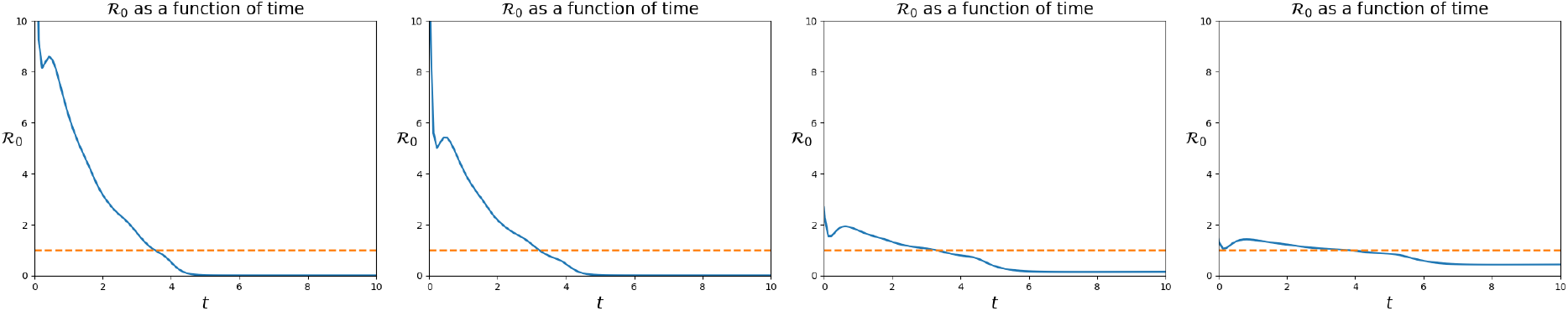
Value of the ℛ_0_ index, as defined in (2.5), as a function of time, according to (2.8), in the four cases, from left to right: *κ* = 0, *κ* as in (3.3), 10 *κ* and 20 *κ*. The effect of quarantine is evident: increasing values of *κ* result in longer periods where ℛ_0_ exceeds one, but with lower values of the index. This typically results in a more bearable stress on the health system.

### 3.4 Residential Care Homes

A recurrent problem in several countries has been the propagation of Covid-19 in care homes. Here, we simulate this phenomenon, showing that the presence of a less controlled area, though containing only one age segment, not only directly suffers from the pandemic, but may well accelerate the virus propagation in the care homes’ neighborhoods.

To this aim, we now integrate (2.8) with the following initial datum:

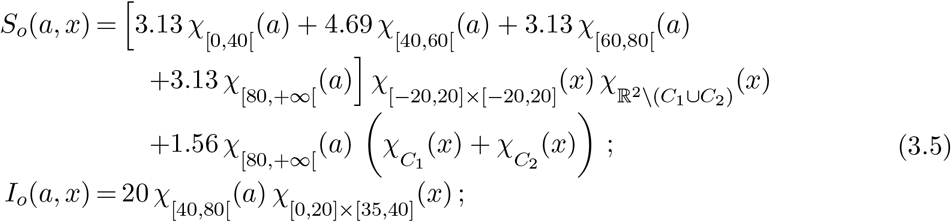

*H_o_*(*a, x*)=0;

*R_o_*(*a, x*)=0.

where the two care homes *C*_1_ and *C*_2_ are located at

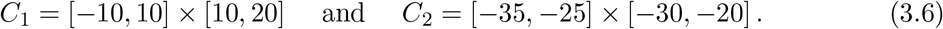

In these regions, only one age class, namely the oldest one, is present and, mostly, less protective measures are adopted. We describe this underestimation of the dangers related to the virus through the function *ρ*:

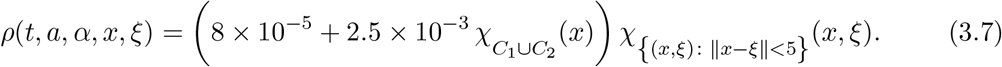

Note that in *C*_1_ and *C*_2_, *ρ* is about 30 times larger than outside these regions.

At time *t* = 0, the *S* population is (approximately) uniformly distributed in [−40, 40] × [−40, 40]. In *C*_1_ and in *C*_2_ only members of the oldest age group (i.e., *a >* 80) are present, see Figure 3.4. Quickly, at time *t* = 3, the virus reaches the first care home *C*_1_, see Figure 9.

**Figure 8:**
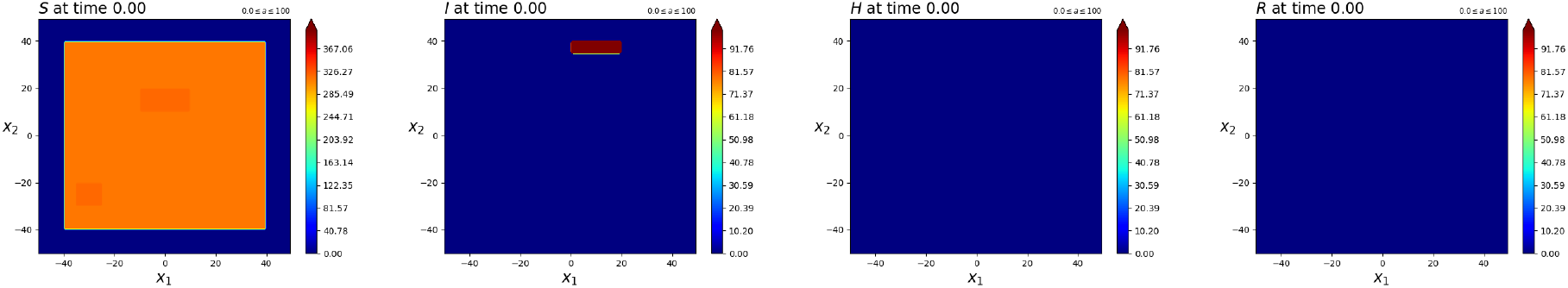
Contour plots of the initial datum for the *“Care Homes”* integration. Note that the total distribution of the *S* population is approximately uniform.

**Figure 9:**
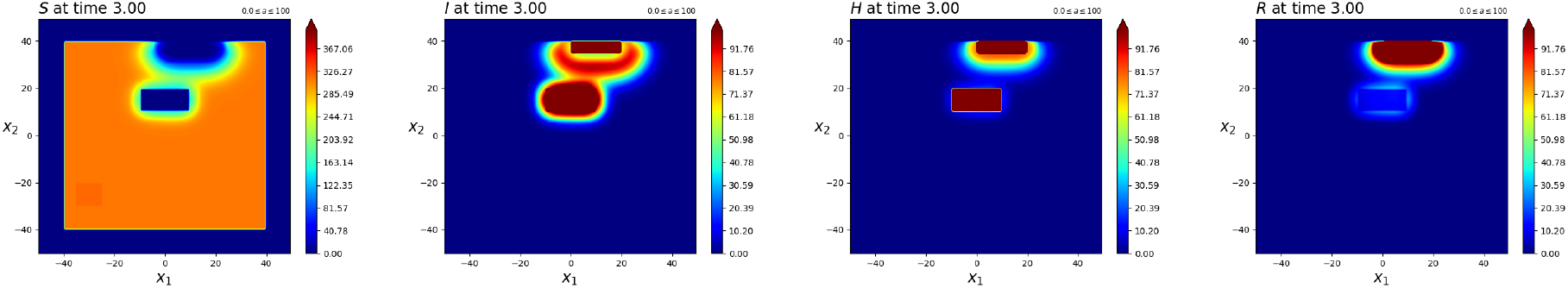
Contour plots of the solution to the *“Care Homes”* integration at time *t* = 3. Note the fast spreading of the disease in *C*_1_ as defined in (3.6). From there, the virus spreads even faster.

As a consequence, the pandemic accelerates and, at time *t* = 7, also *C*_2_ is widely infected, see Figure 10. This further accelerates the spreading, with *C*_2_ clearly acting as a further source of infection, see Figure 11. At time *t* = 10, the two fronts of the pandemic propagation are evident: the first one due to the initial presence of infected individuals, the second one emanating from *C*_2_. Finally the situation at time *t* = 13 is plotted inFigure 12.

**Figure 10:**
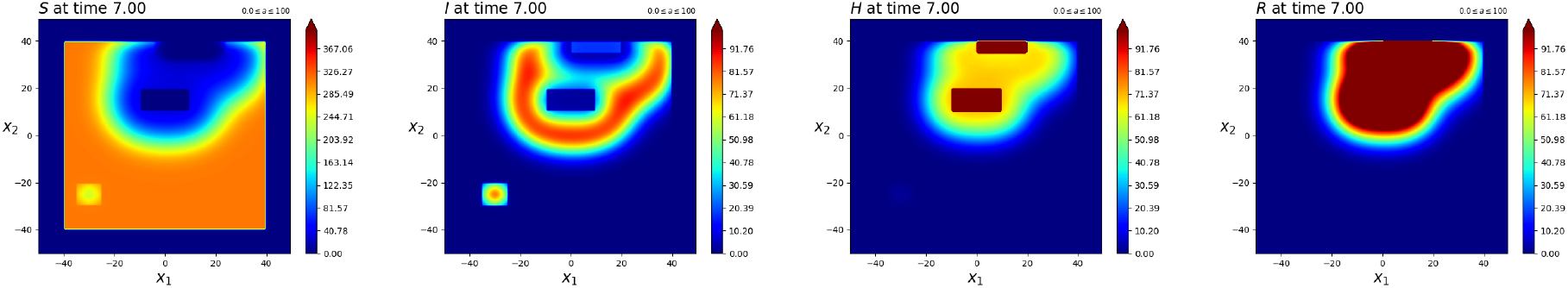
Contour plots of the solution to the *“Care Homes”* integration at time *t* = 7. Note that the care house *C*_2_, as defined in (3.6), is reached by the virus through a very small amount of *I* individuals, so small that it is not highlighted with the current scale. Indeed, contrary to the impression suggested by these figures, the model (2.8) does not allow for any propagation at a distance greater than *δ* = 5, as specified in (3.7).

**Figure 11:**
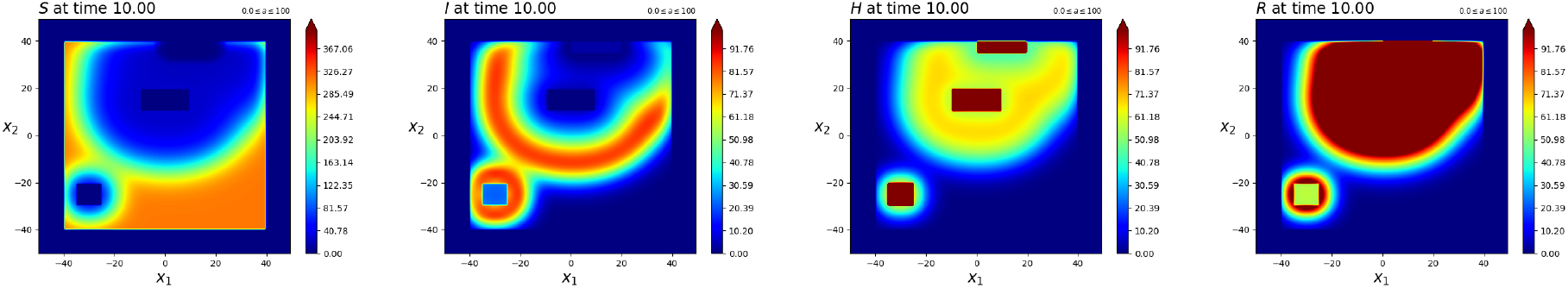
Contour plots of the solution to the *“Care Homes”* integration at time *t* = 10. The front of the pandemic clearly spreads from the top right towards the bottom left of the domain and now the care house *C*_2_, as defined in (3.6), acts as an epidemic outbreak, opening a second front and accelerating the pandemic in the lower left part of the domain.

**Figure 12:**
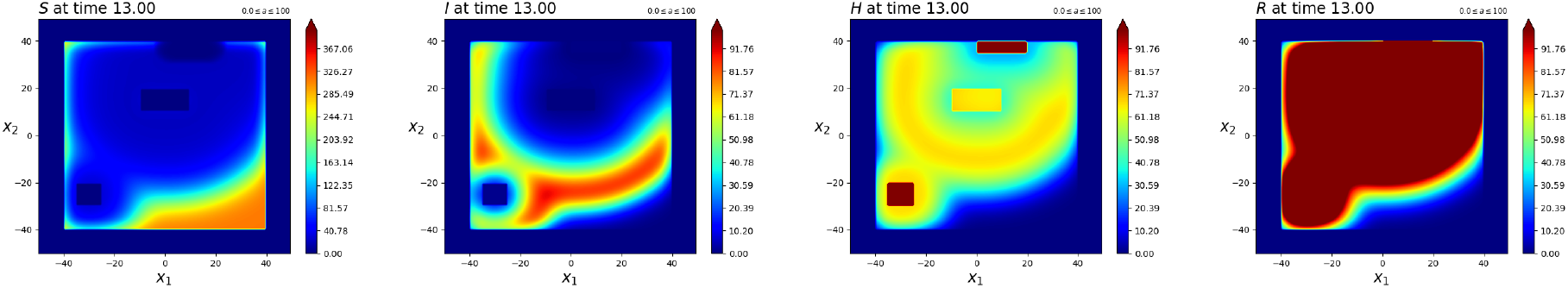
Contour plots of the solution to the *“Care Homes”* integration at time *t* = 13. The front of the pandemic for the *I* population spreads from the care house *C*_2_, as defined in (3.6).

In Figure 13, we see the total amounts of individuals of all ages and over all the domain. Note, in particular, that the initial trend of the *I* population is towards a *decrease*. Nevertheless, the outbreak of the pandemic in the first care home *C*_1_ is able to invert this trend and the total number of infected individuals starts to grow.

**Figure 13:**
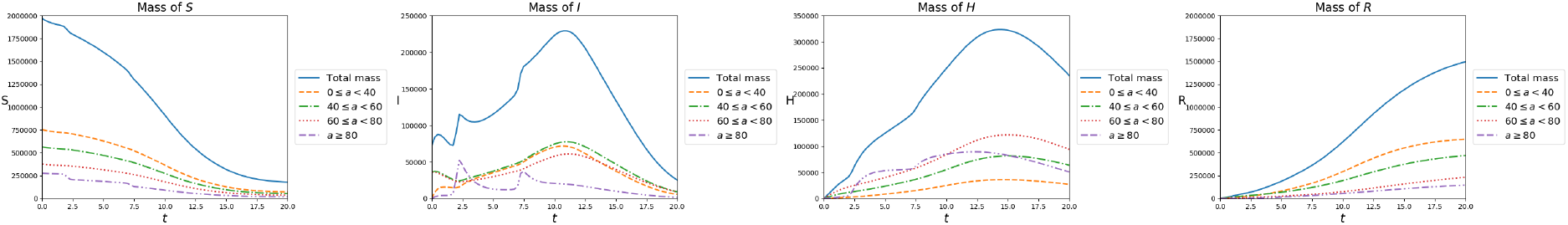
Total amount of individuals of the four populations over all the domain and all age classes, as a function of time, with reference to the *“Care Homes”* integration. Remarkably, the initial tendency of the *I* population is towards a reduction but, as soon as the virus reaches *C*_1_, at about *t* = 2, this tendency is reverted and the pandemic accelerates.

Finally, remark that although the two care homes are rather small with respect to the whole domain, the spreading of the virus in *C*_1_ and in *C*_2_ is very clearly caught by the indexed ℛ_0_ and 𝒬_0_ defined in (2.5), see Figure 14.

**Figure 14:**
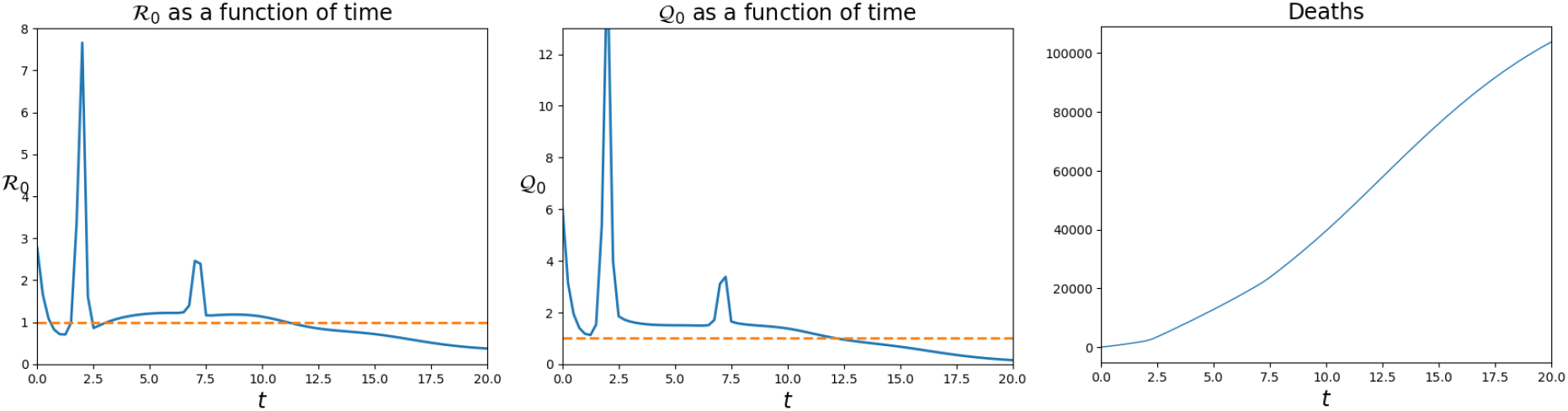
ℛ_0_, left, and Q0, middle, indexes as a function of time in the simulation of the *“Care Homes”* case. The epidemic outbreaks in *C*_1_ and *C*_2_ are clearly visible. Right, the total number of deaths due to the pandemic, as a function of time.

## Data Availability

The sources of the data used are listed in the references with all details.

## Acknowledgments

The authors were partly supported by the GNAMPA 2020 project *”From Wellposedness to Game Theory in Conservation Laws”*. The *IBM Power Systems Academic Initiative* substantially contributed to the numerical integrations.

## Footnotes

1 As usual, if *A* ⊆ R^*N*^, *χA*(*x*) = 1 iff *x* ∈ *A* and *χA*(*x*) = 0 iff *x* ∈ ℛ^*N*^ \ *A*.

